# Fc-gamma receptor expression and cytokine response to intravenous human immunoglobulin in mothers and neonates

**DOI:** 10.1101/2021.09.28.21264275

**Authors:** Stephania Vazquez-Rodriguez, Lourdes A. Arriaga-Pizano, Ismael Mancilla-Herrera, Jessica Prieto-Chávez, Roberto Arizmendi-Villanueva, Ana Flisser, Ethel García-Latorre, Arturo Cérbulo-Vázquez

## Abstract

**Objective:** The use of intravenous human immunoglobulin (IVIg) as adjuvant therapy for sepsis has been shown efficacious in adults, but its use in pregnant women and newborns is controversial. Fc gamma receptors (FcγRs) support the ability of IVIg to stimulate the synthesis of inflammatory mediators and promote phagocytosis by leukocytes, however, the FcγRs expression is differential between adults and neonates. We aimed to explore the effect of IVIg in monocytes and neutrophils from mother and neonates in whole blood cultures.

**Study design:** Whole blood from adults, maternal, and neonates were incubated with LPS and/or IVIg. After 0, 24, and 48 hours, we measured the expression of FcγRs (CD16, CD32, and CD64) and bacterial phagocytosis by monocytes and neutrophils. Also, the concentration of pro-inflammatory cytokines/chemokines was determined.

**Results:** FcγRs expression is quite similar among groups, and the LPS or IVIg challenge did not change the FcγRs expression on monocytes and neutrophils. Also, the LPS or IVIg challenge did not modify phagocytosis capacity in any group. However, IVIg induces a higher IL-8 response in neonates than in adults.

**Conclusion:** Our results suggest that the IL-8 response to IVIg in whole blood from neonates is not dependent on differential FcγR expression.

**Key messages:**

- IVIg challenge in neonates or adults does not induce FcγR change expression on monocytes or neutrophils
- IVIg induces higher IL-8 response in neonates than in adults

## Introduction

Human IVIg is a pharmaceutical preparation based on polyclonal serum, it is used as a substitute in immunodeficiency, and as immunomodulator treatment in autoimmunity or infection diseases (1). Along with antibiotic therapy, IVIg has been used in adults with sepsis (2-4), and has been proposed in neonatal sepsis, however, contradictory results have been reported in neonates. While some do not recommend the use of IVIg in neonates, since it did not improve the clinical condition (5), other studies reports the prevention of early sepsis by IVIg treatment (6). IVIg has been used to treatment of obstetrics indications, such as fetal-neonatal alloimmune thrombocytopenia, antiphospholipid syndrome, and recurrent pregnancy loss (7, 8), however, the effects of IVIg on pregnant women and newborns at the moment of birth are poorly analyzed. The pregnant women and her fetus are sharing multiple mechanisms to regulate the immune system and support the “Immunological Tolerance in Pregnancy” (9, 10), also, the time of birth is an crucial moment where cellular and inflammatory response are necessary for physiologic birth and could be affected by the treatment with IVIg.

Immunomodulatory effects of IVIg are supported by several cellular mechanisms. These include the inhibition of lymphocyte proliferative responses, limitation of inflammatory cytokine response, or the induction of apoptosis, among others (11-13). Most of these mechanisms depend on the binding between antibody-crystallizable fraction (Fc) to Fc-gamma receptors (FcγRs), that are express on almost every immune cell (13). Three different types of FcγR have been described, FcγRI (CD64), FcγRII (CD32), and FcγRIII (CD16) (14). CD64 is a high-affinity receptor to monomeric IgG, while CD32 and CD16 are low-affinity receptors to IgG. Constitutively, monocytes express CD64, macrophages and neutrophils express CD32, and neutrophils and NK cells express CD16 (14), and upon binding of IgG to FcγR, leukocytes mediate phagocytosis and cytokines synthesis (13). However, FcγR is differential between adults and neonates. CD16 and CD32 on monocytes are less expressed on adults than in neonates, and the CD64 expression on granulocytes is higher in neonates than adults (15). Also, it has been reported that after IVIg treatment, the FcγR expression changes on effector cells in mice (16, 17).

To explore the effect of IVIg on neonate immune regulation, we used an *ex vivo* model. Whole blood from adults, pregnant women and neonates were stimulated with lipopolysaccharide (LPS), and the effect of IVIg over the expression of FcγRs, phagocytic capacity of monocytes and granulocytes, and the proinflammatory cytokines response were analyzed.

## Patients and methods

### Patients and sample collection

We included three groups: Healthy non-pregnant women (NP, n=18), healthy pregnant women (P, n=15), and healthy newborn (N, n=18). The Research Committee in the Women’s Hospital in Mexico City reviewed and approved this study (Project: HM: INV/2015:2020). After informed consent was signed, six milliliters of peripheral blood from NP or P group patients were obtained by venipuncture. Also, after placenta delivery, six milliliters of umbilical cord blood (UCB) was obtained by arterial umbilical venipuncture. Blood samples were collected using Vacutainer® plastic sodium heparin tubes (Cat. 367876, BD San Jose, CA, USA) and were processed immediately.

### Cell Culture

One milliliter of adult or neonate whole blood was incubated in 24-well culture plates (Cat 13485, Costar, NY, USA), with or without 10 ng/mL of *Escherichia coli* O55: B5 LPS (Cat. L2880, Sigma Aldrich, St. Louis, MO, USA) and 10 mg/mL of IVIg (Cat. 5240IgG6, Kedrigamma, Kedrion Group S.p.a., Italy) for 0, 24 and 48 hours at 37°C with 5% CO_2_.

### Assessment of FcγR surface expression on monocytes and neutrophils

After culture times (0, 24 and 48 h) with or without LPS and/or IVIg, whole blood samples (50 µL) were mixed with antibodies according to the following panel: CD45-PacificOrange (Cat. MHCD4530, Invitrogen, Waltham, MA, USA), CD14-PE/Cy7 (Cat. 301804, BioLegend, San Diego, CA, USA), CD16-APC/Cy7 (Cat. 302018, BioLegend, San Diego, CA, USA), CD32-PE (Cat. 303206, BioLegend, San Diego, CA, USA), and CD64-APC (Cat. 305014, BioLegend, San Diego, CA, USA). Appropriate isotype controls for each antibody were used. After 15 minutes of incubation, erythrolysis was performed using FACS™ Lysing Solution (Cat. 349202, BD, San Jose, CA, USA). The samples were washed twice with PBS 1× (1,500 rpm, 5 minutes, 4°C), and resuspended in 100 µL of PBS. Thirty thousand single leukocytes were acquired in a FACS Aria IIu flow cytometer (BD Biosciences, San José, CA, USA). The FACS files were analyzed with Infinicyt software 1.8 (Cytognos, Salamanca, Spain). Leukocytes were defined as single cells (FSC-A *vs*. FSC-H plot), CD45 positive, and with the usual size and complexity (SSC *vs*. CD45 plot). Monocytes as SSC^mid^FSC^mid^CD45^+^CD14^+^, and neutrophils were gated as SSC^mid^FSC^mid^CD45^+^CD16^+^. Percentage of positive cells and the mean fluorescence intensity (MFI) for CD64, CD32 and CD16 were calculated.

### Phagocytosis assay

After cell culture times, samples were treated according to the manufacturer’s instructions (pHrodo™ Green *E. coli* BioParticle kit, Cat. P35381, Invitrogen, Waltham, MA, USA). Briefly, blood was incubated with opsonized pHrodo-conjugated *E. coli* bioparticles at 37°C for 15 minutes. Also, blood samples were incubated with pHrodo-conjugated *E. coli* bioparticles on ice for 15 minutes. Blood samples were also immunophenotyped for monocytes and neutrophils expressing CD64, CD32 and CD16. Then, erythrocytes were lysed using buffer A and B, followed by centrifugation and washing. Cell were counted and resuspended in wash buffer to be analyzed by a FACS Aria IIu BD cytometer. Nucleated phagocytes were discriminated using SSC and FSC parameters.

### Cytokine quantification

Also, after culture times and according to the manufacturer’s instructions (LEGENDplex human inflammation kit, Cat. 740808, BioLegend, San Diego, CA, USA) the concentration of cytokines (TNF-α, IL-1β, IL-6, IL-10 and IL-12) and chemokines (IL-8, CCL2) were determined in the plasma. Data were acquired with a FACS Aria IIu BD cytometer. Log-transformed data were used to construct standard curves fitted to ten discrete points, using a 4-parameter logistic model. Concentrations were calculated using interpolations of the corresponding reference curves.

### Statistical Analysis

Data analysis was performed using the Prism 7.0 (GraphPad Software, San Diego, CA, USA). Results are expressed as mean±SD. The Kruskal-Wallis test or two-way ANOVA test with Tukey’s multiple comparisons test was calculated as appropriate. An IC 95% was calculated for every test, and *p*<0.05 was considered as statistically significant.

## Results

### Clinical characteristics

Eighteen NP women and fifteen P women included in the study, were similar age (p=0.9, 24±4 and 25±5 years respectively). All P women were in the third trimester of pregnancy (Mean of age:39 weeks, interval of 37-40.3 weeks). Also, eighteen newborns were analyzed (10 males and 8 females). Apgar’s and Sylverman-Anderson’s scores were normal for healthy neonates. Also, birth temperature (mean:37°C, interval 36-38°C), weight (mean:3,114 g interval 2,360-3,630g), length (mean:49cm, interval 45-51cm), cardiac frequency (124±32 beats/min), and respiratory frequency (45±18 breaths/min) were between the normal limits.

### FcγR expression on monocytes and granulocytes

We counted the lowest percentage of monocytes in P (Mean±SD, 2.2±1.2%) than in N group (8.9±4.4%, p=0.0007, Kruskal-Wallis test, IC 95%, p<0.05), and the highest percentage of granulocytes in P (Mean±SD, 69.2±6.1%) than in N group (40.9±10.4%, p=0.0007, Kruskal-Wallis test, IC 95%, p<0.05). The percentage of monocytes and granulocytes in the NP group was similar to monocytes and granulocytes in the P and N group, no statistical difference was found. An index (MFI / %) was calculated to evaluate the expression of CD16, CD32 and CD64 per cell. Table 1 show the FcγR index in monocytes or neutrophils after 0, 24 and 48 hours of culture, with or without stimulus. The N group show the highest CD16 index in monocytes and neutrophils among groups, this just observed at the start of kinetic (p=0.01), then at 24 and 48 hours the index for all groups goes down, and no difference statistically significant was observed. In addition, IVIg or LPS treatments did not induce significant changes in the CD16 at any kinetic point. The FcγR Index for CD32 in monocytes and neutrophils was similar among the groups, and remained unchanged after LPS stimulus, however IVIg stimulus lead to higher expression in neutrophils only in the NP group (Table 1, p=0.006 and p=0.01). The CD64 index in monocytes show some differences among kinetic in NP and P group, but none after IVIg challenge, in contrast, N group did not show difference among time in the kinetic or by IVIg or LPS challenge. Higher CD64 Index was observed among groups at the 24 and 48 hours. Neutrophils did not show the differences that monocytes do.

**Table 1.** Basal index expression of FcγR on monocytes and neutrophils.

| Index | Hou | NP |  |  | p | P |  |  | p | N |  |  | p | 0p´ | 24p´ | 48p´ |
| --- | --- | --- | --- | --- | --- | --- | --- | --- | --- | --- | --- | --- | --- | --- | --- | --- |
|  |  | 0 | 24 | 48 |  | 0 | 24 | 48 |  | 0 | 24 | 48 |  |  |  |  |
| CD16 MIF/% monocytes |  |  |  |  |  |  |  |  |  |  |  |  |  |  |  |  |
| Medium |  | 282 | 47 | 80 | ns | 549 | 342 | 125 | ns | 2004 | 1125 | 361 | <sup>B</sup>**0.01** | <sup>b0</sup>**0.01** | ns | ns |
| IVIg |  | 282 | 862 | 134 | ns | 549 | 296 | 132 | ns | 2004 | 251 | 253 | <sup>A</sup>**0.03**,<br><sup>B</sup>**0.03** | <sup>b0</sup>**0.04** | ns | ns |
| LPS |  | 282 | 124 | 141 | ns | 549 | 416 | 126 | ns | 2004 | 430 | 270 | <sup>A</sup>**0.02**,<br><sup>B</sup>**0.01** | <sup>b0</sup>**0.01** | ns | ns |
| IVIg + LPS |  | 282 | 124 | 96 | ns | 549 | 416 | 99 | ns | 2004 | 430 | 383 | <sup>A</sup>**0.01**,<br><sup>B</sup>**0.01** | <sup>b0</sup>**0.01** | ns | ns |
| CD32 MIF/% monocytes |  |  |  |  |  |  |  |  |  |  |  |  |  |  |  |  |
| Medium |  | 230 | 310 | 349 | ns | 122 | 239 | 200 | ns | 139 | 227 | 344 | ns | ns | ns | ns |
| IVIg |  | 230 | 77 | 345 | <sup>C</sup>**0.02** | 122 | 182 | 202 | ns | 139 | 127 | 201 | ns | ns | ns | ns |
| LPS |  | 230 | 198 | 216 | ns | 122 | 192 | 144 | ns | 139 | 211 | 183 | ns | ns | ns | ns |
| IVIg + LPS |  | 230 | 198 | 197 | ns | 122 | 192 | 160 | ns | 139 | 211 | 172 | ns | ns | ns | ns |
| CD64 MIF/% monocytes |  |  |  |  |  |  |  |  |  |  |  |  |  |  |  |  |
| Medium |  | 232 | 225 | 300 | ns | 208 | 349 | 191 | <sup>A</sup>**0.03**,<br><sup>C</sup>**0.01** | 135 | 181 | 142 | ns | ns | <sup>c24</sup>**0.009** | <sup>b48</sup>**0.004** |
| IVIg |  | 232 | 276 | 281 | ns | 208 | 270 | 204 | ns | 135 | 153 | 116 | ns | ns | ns | <sup>b48</sup>**0.03** |
| LPS |  | 232 | 432 | 399 | <sup>A</sup>**0.03** | 208 | 317 | 197 | ns | 135 | 197 | 166 | ns | ns | <sup>b24</sup>**0.04** | <sup>b48</sup>**0.04** |
| IVIg + LPS |  | 232 | 432 | 399 | <sup>A</sup>**0.02** | 208 | 317 | 250 | ns | 135 | 197 | 133 | ns | ns | <sup>b24</sup>**0.01** | <sup>b48</sup>**0.005** |
| CD16 MIF/% neutrophils |  |  |  |  |  |  |  |  |  |  |  |  |  |  |  |  |
| Medium |  | 386 | 784 | 273 | <sup>C</sup>**0.04** | 432 | 444 | 328 | ns | 525 | 390 | 221 | ns | ns | ns | ns |
| IVIg |  | 386 | 409 | 312 | ns | 432 | 392 | 341 | ns | 525 | 569 | 235 | <sup>B</sup>**0.01**,<br><sup>C</sup>**0.004** | ns | ns | ns |
| LPS |  | 386 | 405 | 248 | ns | 432 | 355 | 307 | ns | 525 | 335 | 234 | <sup>B</sup>**0.003** | ns | ns | ns |
| IVIg + LPS |  | 386 | 405 | 332 | ns | 432 | 355 | 374 | ns | 525 | 335 | 229 | <sup>B</sup>**0.01** | ns | ns | ns |
| CD32 MIF/% neutrophils |  |  |  |  |  |  |  |  |  |  |  |  |  |  |  |  |
| Medium |  | 83 | 129 | 216 | ns | 145 | 88 | 63 | ns | 89 | 80 | 120 | ns | ns | ns | ns |
| IVIg |  | 83 | 111 | 346 | <sup>B</sup>**0.006**,<br><sup>C</sup>**0.01** | 145 | 37 | 43 | ns | 89 | 45 | 42 | ns | ns | ns | <sup>a48</sup>**0.002**,<br><sup>b48</sup>**0.002** |
| LPS |  | 83 | 104 | 143 | ns | 145 | 264 | 75 | ns | 89 | 101 | 90 | ns | ns | ns | ns |
| IVIg + LPS |  | 83 | 104 | 132 | ns | 145 | 264 | 119 | ns | 89 | 101 | 60 | ns | ns | ns | ns |
| CD64 MIF/% neutrophils |  |  |  |  |  |  |  |  |  |  |  |  |  |  |  |  |
| Medium |  | 100 | 133 | 177 | ns | 159 | 148 | 346 | ns | 93 | 144 | 129 | ns | ns | ns | ns |
| IVIg |  | 100 | 141 | 151 | ns | 159 | 184 | 169 | ns | 93 | 115 | 142 | ns | ns | ns | ns |
| LPS |  | 100 | 134 | 123 | ns | 167 | 184 | 246 | ns | 93 | 130 | 137 | ns | ns | ns | ns |
| IVIg + LPS |  | 100 | 134 | 165 | ns | 167 | 184 | 283 | <sup>B</sup>**0.02** | 93 | 130 | 127 | ns | ns | ns | ns |

Data show the mean value for every FcγR Index in monocytes and neutrophils. Differences between times and groups were calculated by Two-way ANOVA test and Tukey multiple comparisons test. IC 95%, p<0.05. *p*: Between times in the group: **A**: 0 vs. 24; **B**: 0 vs. 48; **C**: 24 vs. 48. *p'*: Between groups at the same time. **a0**: NP vs P; **b0**: NP vs N; **c0**: P vs N; **a24**: NP vs P; **b24**: NP vs N; **c24**: P vs N; **a48**: NP vs P; **b48**: NP vs N; **c48**: P vs N.

**Table 2.**
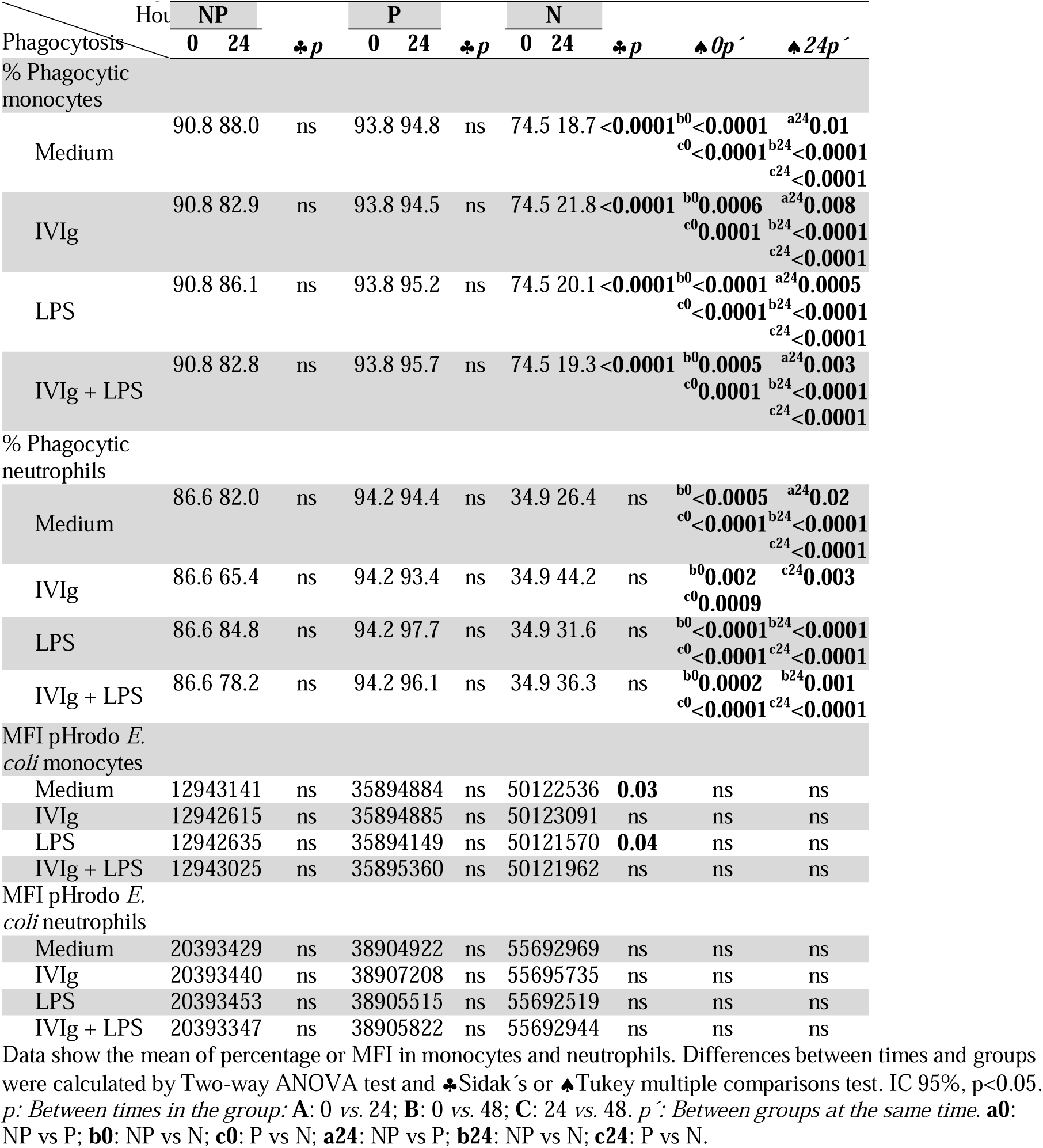
Phagocytosis in monocytes and neutrophils.

### Effect of IVIg on phagocytic capacity

To determine if monocytes and neutrophils modify their ability to engulf and kill pathogens after IVIg/LPS treatments, we exposed previously stimulated whole blood to bacteria and their phagocytic capability was evaluated (endocytosis and bacteria elimination). We observed that monocytes and neutrophils from NP and P groups express similar percentage capacity to internalize bacteria independently of treatment. In contrast, the percentage of phagocytic monocytes in N group was lower than NP or P group (Mean, 74.5 *vs*. 90.8 or 93.8 respectively), this was the case with and without IVIg or LPS (p<0.0001). The lower phagocytic capacity in monocyte’s neonate was shown after 24 hours of culture and *E. coli* challenge for all treatments (p<0.0001). The percentage of phagocytic neutrophils express similar results than in monocytes. In addition, we observed similarities in monocytes and neutrophils capacity to eliminate bacteria from all groups. However, the highest MFI pH for *E. coli* was detected in N group (5,012 for monocytes and 5,569 for neutrophils). Also, only monocyte’s neonate diminish the MFI pHrodo E. coli (bacteria elimination) after 24 hours of challenge with LPS o medium alone, indicating that IVIg could maintain this value in neonates.

### Cytokine response to IVIg

Figure 1 shows the cytokine response to IVIg or LPS challenge. As we expected, in response to LPS, blood cells produce IL-1β, IL-6 and IL-8 in all groups. The magnitude of the response for all cytokines was lower in P women than in NP women and N. In addition, the presence of IVIg alone in adults seemed to have no effect on cytokines production, and it had no additive or inhibitory effects with LPS. In contrast, IVIg alone induced a strong response of IL-8 in Neonates, and it did not affect the response to the LPS challenge.

**Figure 1.**
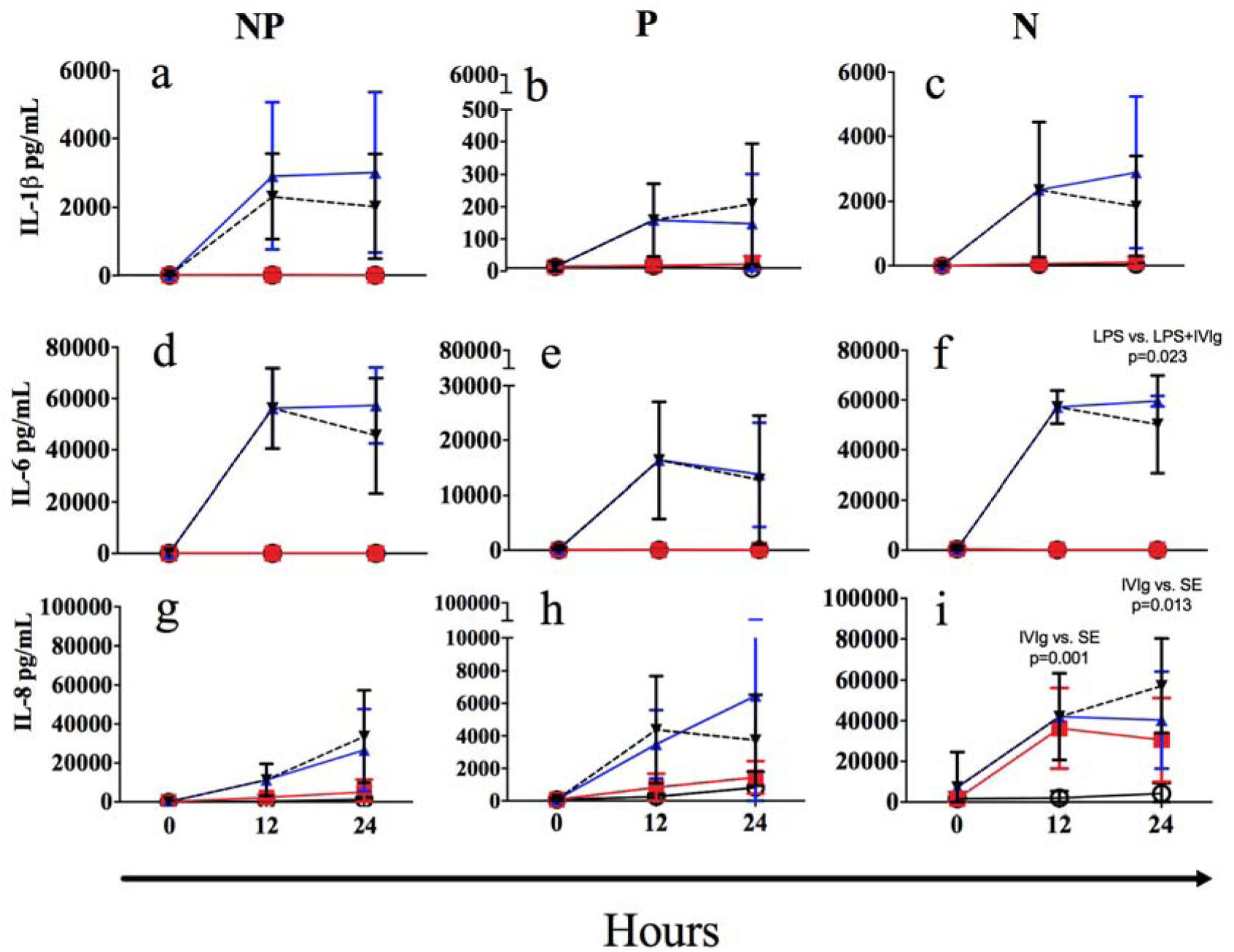
Cytokine response after LPS and IVIg challenge. Whole blood was cultured for 0, 12 and 24 hours in the presence or absence of LPS and IVIg, after each culture time the plasma was collected and cytokine determined. Ten pairs of mothers and neonates are shown. Non-pregnant women (NP, n=10), pregnant woman (P, n=10) and neonate (N, n=10). Empty circle: Medium. Square: IVIg. Triangle Up: LPS, Triangle Down and dash line: LPS + IVIg. Results are expressed as mean±SD. Two-way ANOVA/Tukey’s multiple comparisons test. IC 95%, p<0.05.

## Discussion

Monocytes and neutrophils are fundamental phagocytic cells associated with the inflammatory immune responses (18). They recognize lipopolysaccharide and in response produce large amounts of interleukin (IL)-1β, IL-6, and IL-8 (19-21). IVIg could suppress this inflammatory response in adults but its role in pregnant women and neonates has been rarely studied in a kinetic study. To determine if IVIg can modify the expression of FcγR on monocytes and neutrophils after LPS exposure, we evaluated the kinetics of single or combined stimulation. Also, we evaluated their phagocytic capacity and the cytokine response after IVIg treatment *ex vivo*.

The IgG capacity to stimulate FcγR could explain the biological activities of IVIg. IgG-FcγRs engagement could support pro-inflammatory or anti-inflammatory responses. Our results suggest that the biological activity of IVIg does not depend on CD16, CD32 and CD64 expression on monocytes and neutrophils. On the other hand, CD32b isoform (FcγRIIb) express immunoreceptor tyrosine-based inhibitory (ITIM) motifs, resulting in inhibitory response (14, 22). Also, it has been reported that IVIg infusion increases FcγRIIb expression in myeloid cells, suggesting a possible anti-inflammatory mechanism (3, 11, 13). We did not analyze the percentage of CD32b monocytes or neutrophils, more studies are necessary to solve this question. Maeda *et al*. (15) reported that neonatal granulocytes expressed higher levels of CD64, and Luppi *et al*. (23) and Davis *et al*. (24) reported that CD64 expression in polymorphonuclear cells gradually increased in the third trimester of pregnancy. Our study shows that the expression per cell for CD16, CD32 and CD64 in monocytes and neutrophils are similar, suggesting that FcγRs are not a limitation to possible differential response to IVIg in adults and neonates.

Also, FcγRs participated in the phagocytosis mediated by antibodies. Contrary to Gille *et al*. reports. (25), we did not observe an increase in phagocytosis after IVIg treatment. Gille’s study used isolated mononuclear cells (*in vitro* model), but we used whole blood (*ex vivo* model). Their model explored the contribution of monocytes, meanwhile we tested the combined response of both monocytes and neutrophils. We argue that our *ex vivo* model is closer to the results observed *in vivo*, however, in our study less detail can be observed and the participation of several variables *in vivo* are unknown. We showed that both groups of adults (pregnant and non-pregnant) had more than 80% of monocytes doing phagocytosis of *E. coli*, and was maintained after 24 hours of culture. In contrast, only ≈75% of neonatal monocytes had this ability, and it dropped to ≈20% after 24 hours of culture. This functional limitation in neonates could be due to increase of apoptosis to solve the physiologic leukocytosis in neonates, and limiting some functional capacities such as phagocytosis. Despite this condition, IVIg or LPS stimulus did not change the similar response of phagocytosis in adults and neonates.

IVIg limit the production of inflammatory cytokines such as TNF-α, IL-1β, and IL-6 in patients with sepsis (26-28). However, some studies showed that IVIg *in vivo* could lead to both inhibitory or productive effects for certain interleukins like IL-6 (29, 30). Our results showed that IVIg upregulates IL-8 production in neonates but not in adults, suggesting that the use of IVIg as an immunomodulatory agent may be especially important to modulate IL-8 in neonates. Interestingly, we observed also a low IL-8 response in the mother, suggesting pregnant status is an important condition that could modulate the effects of IVIg. This high response of IL-8 to IVIg could promote the migration of neutrophils that support inflammation in newborns. More studies are necessary to know if this effect of IVIg could support substantial help to medical treatment in neonates.

Our study has some limitations as small sample sizes and short kinetics; however, the study shows differential IL-8 responses among the groups, indicating that IVIg could upregulate different mechanisms in adults and neonates. Our model may not reflect the neonatal response once several days pass after birth, or particular perinatal response when an infection is acquired at birth. More experiments should be conducted to elucidate differences in the responses to IVIg treatment between adults and neonates.

## Data Availability

The data that support the findings of this study are available on request from the corresponding author, A Cerbulo-Vazquez. The data are not publicly available due to [restrictions e.g. their containing information that could compromise the privacy of research participants].

## Acknowledgements

The authors thank the staff at the labor room in Hospital de la Mujer. CONACYT SALUD-2010-C01-141102 funded this work, National Scholarship CONACyT number 289859. The authors want to thank Manuel Flisser for financing the kits to quantify the soluble cytokines: LEGENDplex Human Inflammation and the kits to determine the phagocytic capacity: pHrodo™ Green *E. coli* BioParticle.

## Data availability

The data that support the findings of this study are available on request from the corresponding author, ACV. The data are not publicly available due to [restrictions e.g. their containing information that could compromise the privacy of research participants].

